# Level and determinants of willingness to pay for rapid COVID-19 testing delivered through private retail pharmacies in Kenya

**DOI:** 10.1101/2021.10.10.21264807

**Authors:** Jacob Kazungu, Audrey Mumbi, Precious Kilimo, Jessica Vernon, Edwine Barasa, Peter Mugo

## Abstract

**Introduction:** To support the government response to the coronavirus disease 2019 (COVID-19) pandemic, accessible and sustainable testing approaches are needed. Private retail pharmacies are a key channel through which communities can access COVID-19 testing. We examined the level and determinants of the willingness to pay (WTP) for rapid COVID-19 testing delivered through private retail pharmacies in Kenya.

**Methods:** Data was collected following a cross-sectional double-bounded dichotomous choice contingent valuation survey across 341 clients visiting five private retail pharmacies in Nairobi, Kisumu and Siaya counties.

**Results:** Our findings indicate mean and median WTP levels of KES 611 (US$ 5.59) and KES 506 (US$ 4.63), respectively. Estimated WTP varied across counties and increased with household income and self-reported interest in pharmacy-based COVID-19 rapid testing.

**Conclusion:** These findings can inform price setting, price differentiation, price subsidization and other program design features geared towards enhancing affordability, equity, and uptake.

**Key Questions:** *What is already known?:*

- The Coronavirus disease 2019 (COVID-19) global pandemic continues to cause great morbidity, mortality, social and economic burden.
- Pharmacies in Kenya have been involved in the delivery of several health interventions, such as malaria rapid testing, HIV self-testing, and other disease screening services.
- While COVID-19 testing remains an important response strategy to the ongoing COVID-19 pandemic, it is not clear how much pharmacy clients in Kenya and similar settings would be willing to pay (WTP) to obtain rapid COVID-19 testing at pharmacies

*What are the new findings?:*

- The mean and median willingness to pay (WTP) for a rapid test delivered at a private retail pharmacy was KES 611 (US$ 5.59) and KES 506 (US$ 4.63), respectively.
- WTP varied by county, hence, the need for county-specific price-setting for pharmacy-based COVID-19 testing.
- WTP increased with household income and interest in getting the COVID-19 test at a private retail pharmacy.

*What do the new findings imply?:*

- A better understanding of the user’s willingness to pay price that can guide price setting, price differentiation, price subsidization and other program design features geared towards enhancing affordability, equity, and uptake.

## 1.0 INTRODUCTION

The Coronavirus disease 2019 (COVID-19) global pandemic continues to cause great morbidity, mortality, social and economic burden [1-3]. As of 29^th^ September 2021, a total of 233,633,337 confirmed cases and 4,780,890 deaths had been reported globally [4]. Early detection of infection, a key prevention and management strategy, heavily relies on the presence of adequate testing and diagnostic capacity [5-7]. However, low- and middle-income countries (LMICs) such as Kenya continue to experience low detection capacity. Existing evidence indicates that “for every COVID-19 test conducted in LMICs, over 50 are being conducted in high-income countries” [8].

Detection of COVID-19 in Kenya currently relies mainly on laboratory PCR testing, which is expensive given that it requires highly qualified staff and specialized equipment. Results of PCR testing can be obtained after 24-48 hours, creating a potential delay in prevention and management decision-making. A recent analysis in Kenya indicated that it costs KES 1,816 (US$ 17.22) per COVID-19 test [9], a price that may not allow mass testing or adequate coverage of testing required for an effective pandemic response.

Rapid diagnostic tests (RDTs) have been developed to support the early detection of COVID-19 infection. The first RDT device for the diagnosis of COVID-19 entered the market at the end of March 2020, and as of 27th September 2021, there were a total of 354 antigen and 260 antibody RDTs that are commercially available or in development. However, very few of these tests have been developed in Africa or are in use in LMICs [10].

Private retail pharmacies (also referred to as chemists or community pharmacies) present a unique setting for rapid COVID-19 testing given that they are often the first or only point of contact of patients with the healthcare system [11, 12]. The reasons for preferential care-seeking at pharmacies vary, but the main ones are greater accessibility (pharmacies are more numerous, more conveniently located, and provide speedier services), lower cost, and greater perceived privacy, compared to health facilities [13, 14]. Recent evidence points to a decline in the utilisation of inpatient and outpatient services in Kenya [15]; pharmacies could be absorbing some of this patient traffic. Pharmacies in Kenya have been involved in the delivery of several health interventions, such as malaria rapid testing, HIV self-testing, and other disease screening services [16-20]. Consequently, private retail pharmacies can be used to deliver COVID-19 rapid testing services, hence expanding testing coverage and access to affordable and timely testing.

However, it is not clear how much pharmacy clients in Kenya and similar settings would be willing to pay (WTP) to obtain rapid COVID-19 testing at pharmacies. Although several WTP studies have been conducted, these have so far focused on other diseases such as HIV or vaccines or conducted in high-income countries [21-25]. Against this backdrop, this study used a contingent valuation method (CVM) to examine the level and determinants of the willingness to pay for rapid COVID-19 testing delivered through private retail pharmacies in Kenya.

## 2.0 METHODS

### 2.1 Study design and setting

This study involved a cross-sectional survey conducted at five private retail pharmacies in Nairobi, Kisumu and Siaya counties of Kenya. As of 2020, the three counties hosted 32% (1,602/5,033) of licensed pharmacies in the country [26] and, as of 17^th^ April 2021, contributed 55% (83,607/152,523) of reported COVID-19 cases [27, 28]. Study pharmacies were randomly selected from a list maintained by Maisha Meds, a health technology and data analytics organization and a collaborator on this study. Maisha Meds has built a network of private pharmacies and clinics in Kenya, Uganda, and Tanzania, through a free Android™ point-of-sale application that pharmacies use to manage inventory and track sales; re-order discounted medicines from pre-qualified suppliers and receive payments from clients. Nairobi county was included in the study because it is the main base county for the investigators, has the second-highest number of private retail pharmacies within the Maisha Meds network and has had the highest number of reported COVID-19 cases in Kenya. On the other hand, Kisumu County was included as it has the highest number of pharmacies in the Maisha Meds Network and is the main base for the Maisha Meds field team. Lastly, Siaya county was included to primarily represent a more rural setting compared to Nairobi and Kisumu and also hosts the third-largest number of pharmacies within the network (Table 1).

**Table 1:** Distribution of pharmacies in the Maisha Meds Network

| Region | County | Number of pharmacies | Sub-total | Proportion |
| --- | --- | --- | --- | --- |
| Coast | Mombasa | 4 | 8 | 3% |
| Coast | Kwale | 3 |  |  |
| Coast | Kilifi | 1 |  |  |
| Nairobi and Central | Nairobi | 33 | 48 | 18% |
| Nairobi and Central | Machakos | 5 |  |  |
| Nairobi and Central | Kiambu | 5 |  |  |
| Nairobi and Central | Embu | 1 |  |  |
| Nairobi and Central | Tharaka-Nithi | 1 |  |  |
| Nairobi and Central | Nyeri | 1 |  |  |
| Nairobi and Central | Murang'a | 1 |  |  |
| Nairobi and Central | Kirinyaga | 1 |  |  |
| Rift valley | Nakuru | 7 | 23 | 9% |
| Rift valley | Kericho | 5 |  |  |
| Rift valley | Uasin Gishu | 3 |  |  |
| Rift valley | Kajiado | 3 |  |  |
| Rift valley | Nandi | 2 |  |  |
| Rift valley | Bomet | 2 |  |  |
| Rift valley | Baringo | 1 |  |  |
| Western | Kisumu | 79 | 190 | 71% |
| Western | Siaya | 24 |  |  |
| Western | Migori | 21 |  |  |
| Western | Homa Bay | 16 |  |  |
| Western | Kakamega | 13 |  |  |
| Western | Bungoma | 12 |  |  |
| Western | Busia | 11 |  |  |
| Western | Vihiga | 9 |  |  |
| Western | Nyamira | 2 |  |  |
| Western | Kisii | 2 |  |  |
| Western | Trans-Nzoia | 1 |  |  |
| **TOTAL** | **29** |  | **269** | **100%** |

### 2.2 Study population and Sampling

The study targeted adults (18+ years) seeking any service at a study pharmacy and willing to provide written informed consent to participate in the study. While there is no consensus on sample size determination in contingent valuation methods [29, 30], contingent valuation studies in the health sector have used a sample size of 300 on average. The study targeted a minimum sample size of 300 clients. The clients were selected through a systematic random sampling approach where every second eligible client visiting a study pharmacy was invited to participate in the study.

### 2.3 Data collection

We conducted a willingness to pay survey using the contingent valuation method. Contingent valuation is a survey-based approach for eliciting the values people attach to goods or services by asking them directly, even if they do not currently use the goods or services themselves [31]. Data was collected following a double-bounded dichotomous choice approach as it has been shown to increase the statistical efficiency in estimation and generates more data that can better inform price-setting, compared to single-bounded approaches [32-34].

Data was collected by trained research assistants using tablets with the questionnaire programmed into REDCap. The questionnaire captured participants socio-demographic characteristics (age, gender, household monthly income, level of education, marital status, employment status, and county of residence), the WTP questions and participant’s attitudes towards pharmacy-based COVID-19 testing (coping mechanism when the prices of the COVID-19 testing kit was above their WTP level and their interest in pharmacy-based COVID-19 testing) (Appendix 1).

For the WTP questions, participants were asked to indicate whether they would be willing to pay (Yes or No) the price of the COVID-19 test kit at a private retail pharmacy at a given bid (KSh 500 or KSh 1000). Participants were randomly assigned to the two starting bids using the randomization module in REDCap [35].

In line with the double-bounded dichotomous choice contingent valuation format, respondents were then presented with a second bid indicating a higher value (1000 or 1500, respectively) when the response to the first bid was “Yes” and a lower bid (300 or 750, respectively) when the answer to the first bid was “No”. These bid amounts were considered realistic prices considering the likely prices of the kits from manufacturers and therefore avoiding the starting point bias as highlighted in a previous study [36].

In addition to the second question, all respondents were asked to state the maximum amount they would be willing to pay to get a COVID-19 test at a pharmacy. Prior to the final survey, a pilot study with a sample of 31 respondents was done to assess whether participants understood the questions and to explore areas for refining the questionnaire. **Figure 1** shows the flow of questions of the WTP section of the questionnaire for the two blocks.

**Figure 1:**
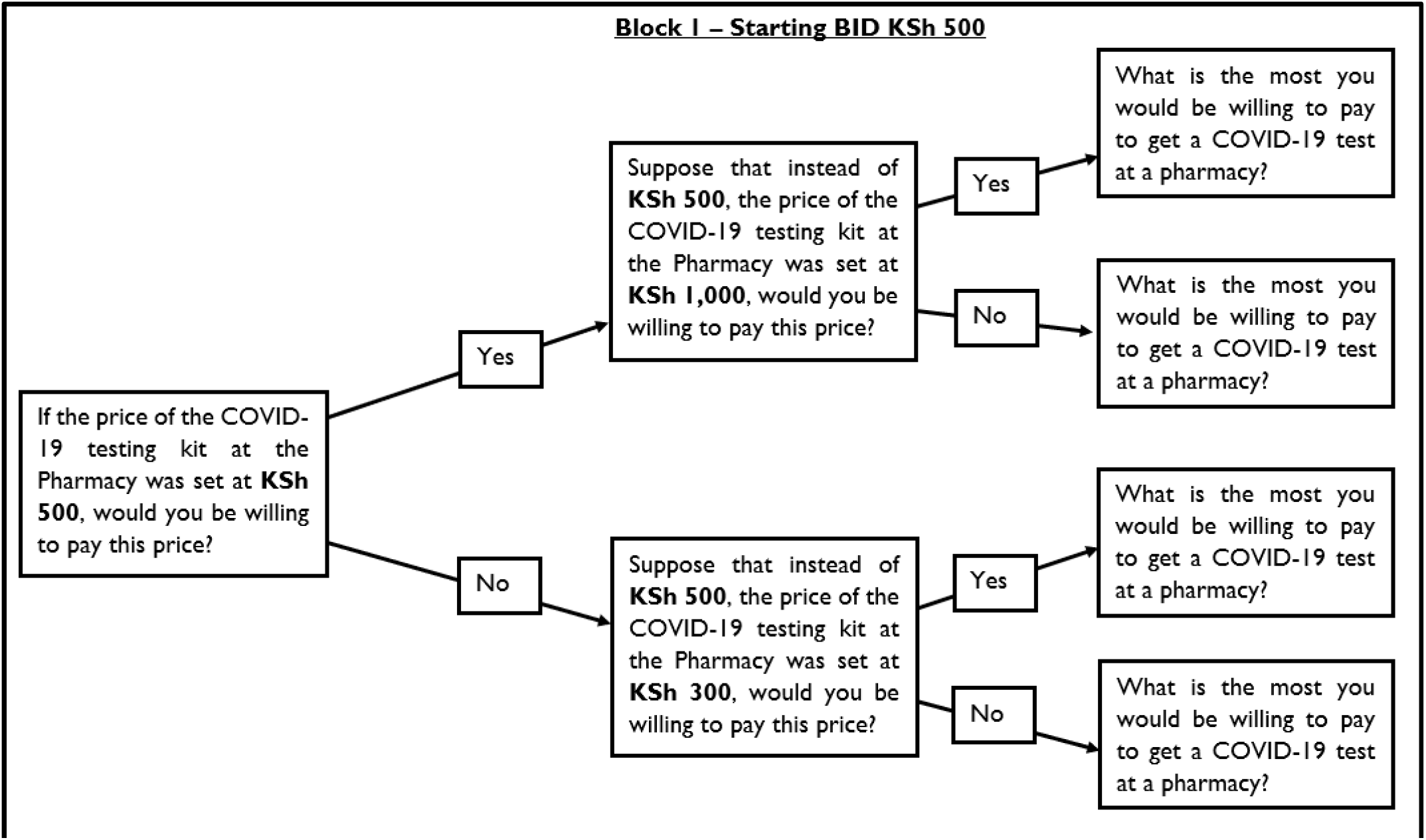
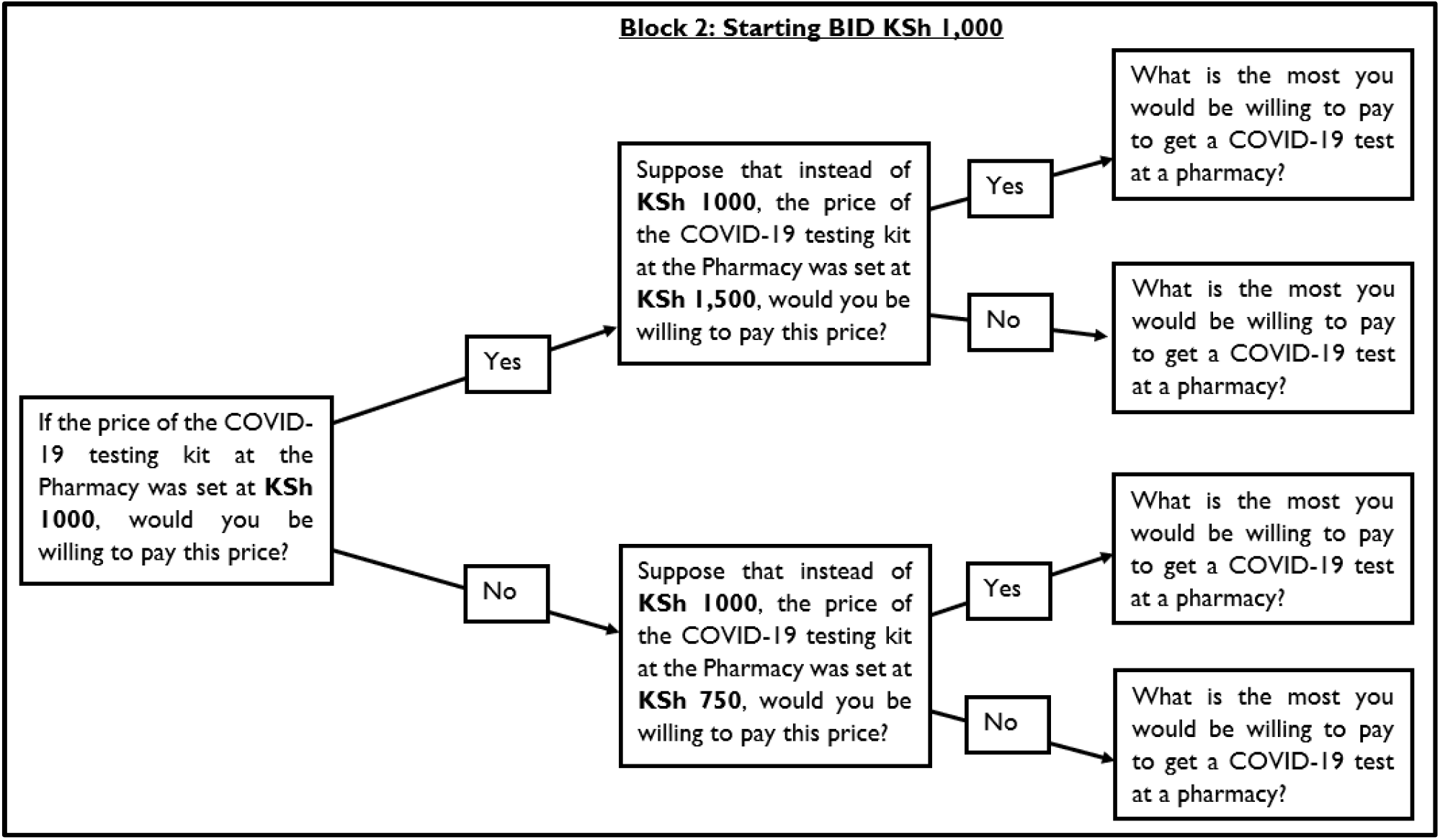
Flow charts of the WTP section of the questionnaire for block 1 with a starting bid of KES 500 and block 2 with a starting bid of KES 1,000.

### 2.4 Data Analysis

Four types of analyses were conducted. First, we conducted descriptive analyses including means and percentages to present the distribution of the study sample and the maximum WTP price across selected sociodemographic factors. Second, we fitted a log-logistic econometric model in R version 4.0.3 following the DCChoice package [31, 37]. Given the structure of the data in double-bounded dichotomous choice contingent valuations, there were four possible outcomes to the responses:

1. YY: yes to the first bid, and yes to the second one; Meaning that a respondent’s WTP > KES 1000 or KES 1,500 (for starting bids of KES 500 or KES 1000 respectively)
2. YN: yes to the first bid, and no to the second one; Meaning that the respondent’s WTP lies between – KES 500 | KES 1,000 ≤ WTP ≤ KES 1,000 | KES 1,500
3. NY: no to the first bid, and yes to the second one; Means KES 300 | KES 750 ≤ WTP ≤ KES 500 | KES 1,000
4. NN: no to the first bid, and no to the second one; Means WTP < KES 300 | KES 750

Given these outcome options, the model can be expressed as:

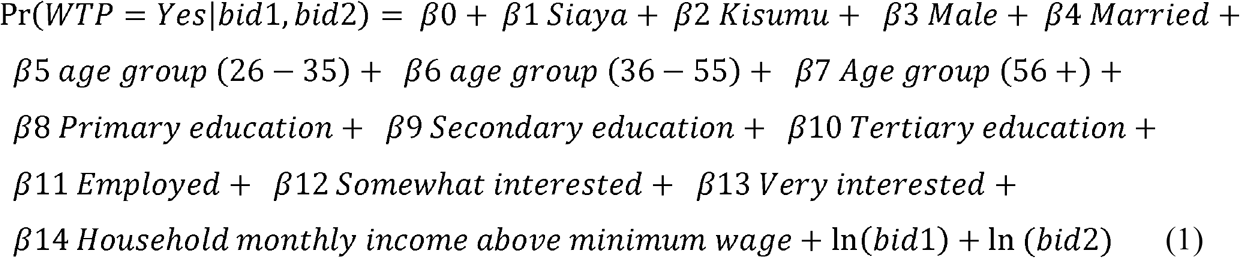

In R, the model can be represented as:

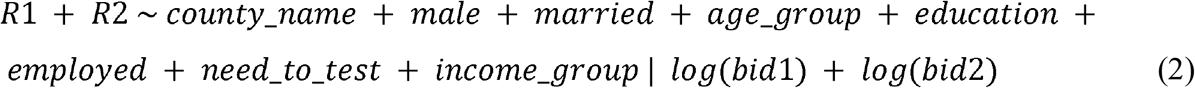

where *bid1* and *bid2* and *R1* and *R2* are the prices (in KES) and responses (Yes or No) to the first question and second questions respectively.

Based on the Hanemann model [38], when the WTP is non-negative, the mean WTP was estimated as below:

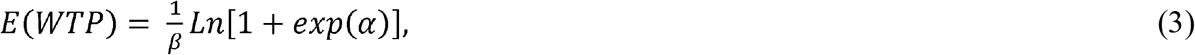

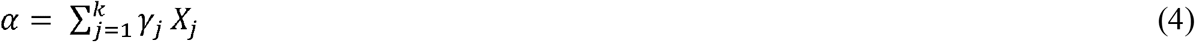

Where β is the coefficient of the bid value; a is the estimated constant term or the macro constant term, which is the product of the regression coefficients for the constant term and other independent variables and their mean values. X is the other socioeconomic variable for the respondent, and g is the corresponding coefficient. In this analysis, we present the mean WTP price (truncated at the maximum bid) and the median WTP.

Third, we plotted demand curves and the price elasticity of demand (PED) by county and overall. The demand curves presented a linear relationship indicating the proportion of the clients willing to buy COVID-19 test kits (X-axis) against the respective WTP prices (Y-axis). Based on these curves, we then computed the PED for each of the counties and an overall estimate for all counties. PED was computed to show how sensitive demand is to changes in price [39] across the three counties. Demand is said to be price elastic when the absolute PED value is greater than 1 and price inelastic when the PED value is between 0 and 1. PED was calculated as below:

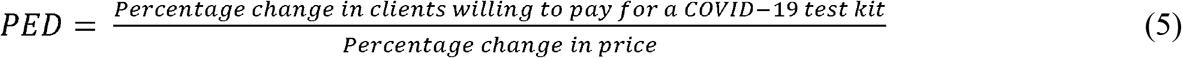

Fourth, we explored the stated maximum WTP price using means, median and fitted a probit model. We computed the mean and median maximum WTP prices stated by respondents across the name of the county, and respondents sociodemographic factors including age, gender, marital status, level of education, household income group and the respondent’s level of interest in the COVID-19 test at a pharmacy if in need. We examined differences in the means of the maximum WTP prices across selected sociodemographic factors using unpaired student T-tests [40]. Besides, we fitted a Tobit regression model to estimate the determinants of the maximum amount pharmacy clients’ WTP for COVID-19 testing at a private retail pharmacy. The Tobit model was considered appropriate for this analysis as the data is zero-censored where individuals could not state a maximum WTP price of less than zero [41]. The linear function of the relationship between the respondent’s maximum WTP price and sociodemographic factors was given by:

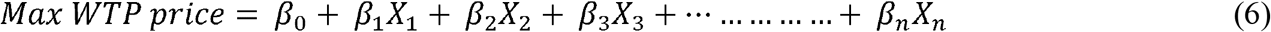

Where: β is a vector of the coefficients and X represents the sociodemographic factors.

We used an exchange rate of US$1=109.32 Kenya shillings (KES), derived from oanda.com accessed on 6^th^ October 2021, to convert KES to US$. We report our findings in KES and US$.

### 2.5 Ethics statement

We received ethical approval from the Kenya Medical Research Institute (KEMRI) Scientific and Ethics Review Unit (SERU) under SERU number SSC: 4115. Additionally, the study was permitted by the National Commission for Science, Technology and Innovation (NACOSTI) as well as the Council of Governors. We also obtained written informed consents from all participants who agreed to participate in the study.

## 3.0 RESULTS

### 3.1 Sample characteristics

**Table 2** shows the distribution of the study sample characteristics for the WTP survey. Overall, the survey included 341 respondents across the three counties. Most respondents were male (55%), married (68%), employed (68%) and had a secondary or higher level of education (72%). over 90% of respondents indicated some interest in getting a COVID-19 test at a private retail pharmacy if they needed to get tested.

**Table 2:** Participant characteristics.

|  | **Total sample n (%)** |
| --- | --- |
| **All respondents across all counties** | 341 (100%) |
| **Survey Block** |  |
| Block 1 | 173 (51%) |
| Block 2 | 168 (49%) |
| **County** |  |
| Nairobi | 117 (34%) |
| Siaya | 81 (24%) |
| Kisumu | 143 (42%) |
| **Gender** |  |
| Female | 154 (45%) |
| Male | 187 (55%) |
| **Current marital status** |  |
| Not married | 110 (32%) |
| Married | 231 (68%) |
| **Age group (years)** |  |
| 18-25 | 87 (25%) |
| 26-35 | 125 (37%) |
| 36-55 | 96 (28%) |
| 56+ | 33 (10%) |
| **Currently employed** |  |
| No | 108 (32%) |
| Yes | 232 (68%) |
| **Level of education** |  |
| No Education | 14 (4%) |
| Primary education | 81 (24%) |
| Secondary education | 116 (34%) |
| Tertiary education | 130 (38%) |
| **Household Income group\*\*** |  |
| Below monthly minimum wage | 177 (52%) |
| Above monthly minimum wage | 164 (48%) |
| **Interest in COVID-19 test at Pharmacy if in need** |  |
| Not at all interested | 14 (4%) |
| Somewhat interested | 73 (21%) |
| Very interested | 254 (75%) |

\*\* The current minimum wage for an individual in Kenya is KES 13,572

### 3.2 Willingness to Pay levels for rapid COVID-19 testing

Overall, the mean WTP was KES 610.72 (US$ 5.59) (95% confidence interval (CI): 481.19 – 665.55) for one COVID-19 rapid test at a private retail pharmacy. The median WTP was KES 505.84 (US$ 4.63) (95% CI: 385.02 – 572.20). **Table 3** presents the results from the econometric model. Of the sociodemographic variables controlled for, respondents from Kisumu, having some interest in getting a COVID-19 rapid test at a pharmacy and household income were the significant factors associated with WTP. Specifically, clients from Kisumu were willing to pay a lower price compared to their counterparts from Nairobi (p<0.001). Those who were somewhat (p=0.014) or very interested (p = 0.002) in getting tested at a pharmacy were willing to pay a higher price. As expected, individuals from households with a monthly income above the minimum wage were significantly p and positively associated with their WTP (p<0.001).

**Table 3:** Log-logistic regression results of the willingness to pay for COVID-19 testing at private retail pharmacies in Kenya.

|  | Coefficient | 95% Confidence Intervals |  | P-value |
| --- | --- | --- | --- | --- |
|  |  | Lower | Upper |  |
| **Intercept** | **11.76** | **8.76** | **14.76** | **<0.001** |
| **Log (BID)** | **-2.36** | **-2.70** | **-2.02** | **<0.001** |
| **County (Ref. Nairobi)** |  |  |  |  |
| Siaya | 0.35 | -0.53 | 1.23 | 0.436 |
| Kisumu | **-1.44** | **-2.01** | **-0.87** | **<0.001** |
| **Gender (Ref Female)** |  |  |  |  |
| Male | 0.45 | -0.01 | 0.91 | 0.057 |
| **Current marital status (Ref. Not Married)** |  |  |  |  |
| Married | -0.29 | -0.79 | 0.21 | 0.260 |
| **Age group (Ref. 18-25 years)** |  |  |  |  |
| 26-35 | 0.17 | -0.43 | 0.76 | 0.583 |
| 36-55 | 0.19 | -0.47 | 0.84 | 0.576 |
| 56+ | -0.38 | -1.31 | 0.56 | 0.432 |
| **Level of education (Ref. No Education)** |  |  |  |  |
| Primary education | 0.52 | -0.81 | 1.85 | 0.442 |
| Secondary education | 0.19 | -1.17 | 1.54 | 0.786 |
| Tertiary education | 0.99 | -0.38 | 2.36 | 0.158 |
| **Employment status (Ref. Not employed)** |  |  |  |  |
| Employed | -0.32 | -1.04 | 0.40 | 0.383 |
| **Interest in COVID-19 test at Pharmacy if in need (Ref. Not at all interested)** |  |  |  |  |
| Somewhat interested | **2.12** | **0.43** | **3.81** | **0.014** |
| Very interested | **2.67** | **1.00** | **4.34** | **0.002** |
| **Household income group (Ref. Below monthly min. wage)** |  |  |  |  |
| Above monthly minimum wage | **1.09** | **0.49** | **1.69** | **<0.001** |
| **Number of Observations** | 340 |  |  |  |
| **Log-likelihood** | -359.93 |  |  |  |
| **Likelihood Ratio test** | 120.60 |  |  |  |
| **AIC** | 751.86 |  |  |  |
| **BIC** | 813.12 |  |  |  |
| **P-value** | <0.001 |  |  |  |

### 3.3 Demand for rapid COVID-19 testing and price elasticity of demand (PED)

As anticipated, the proportion of clients willing to pay for rapid COVID-19 testing at private retail pharmacies reduced as the prices increased in line with the demand theory (**Figure 2**). For instance, when all counties are considered, 53.2% of the clients would be willing to pay KES 500 (US$ 4.57), 42.4% would be willing to pay KES 750 (US$ 6.86), 31.6% would be willing to pay KES 1,000 (US$ 9.15) and only 9.9% would be willing to pay KES 1,500 (US$ 13.75).

**Figure 2:**
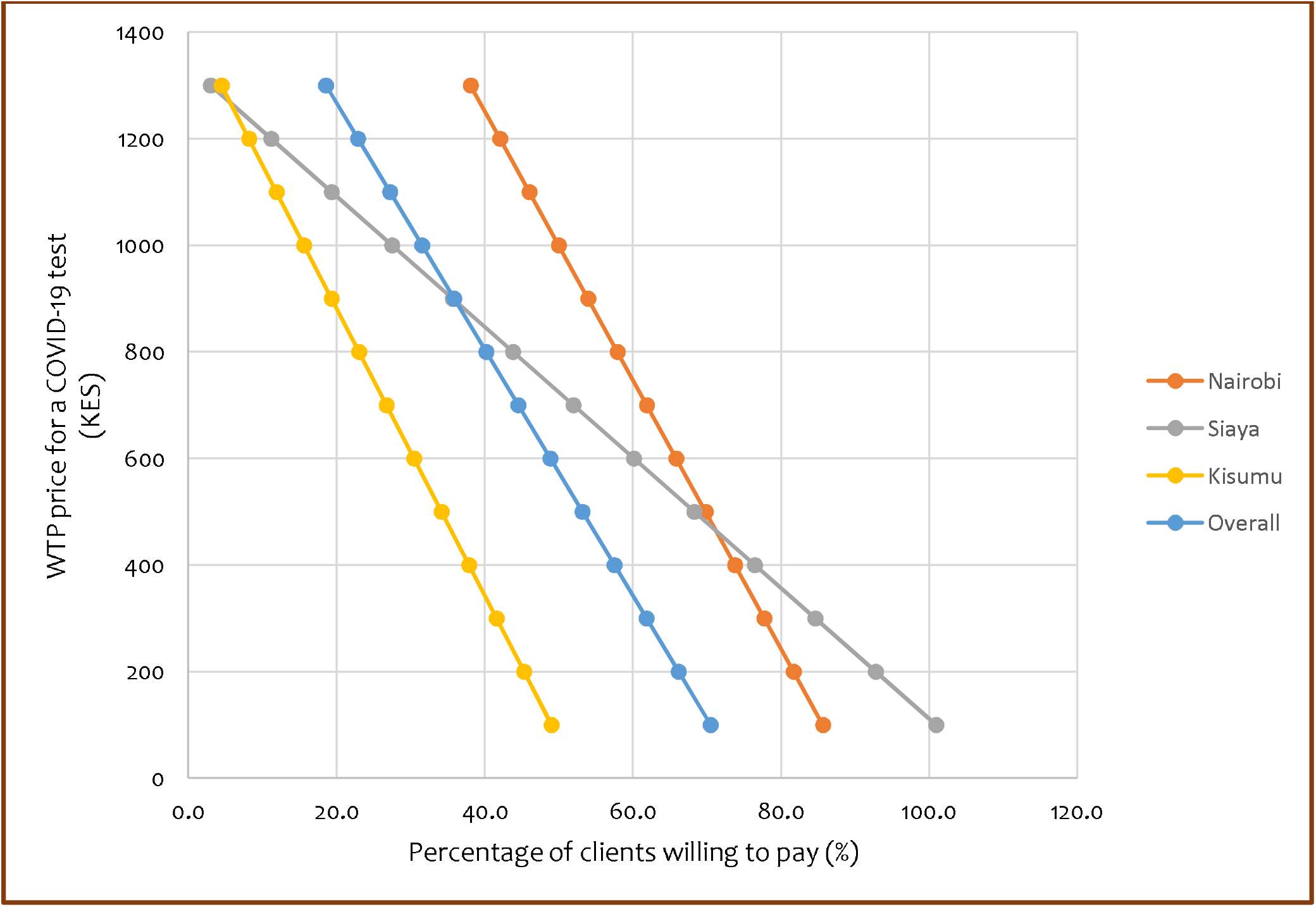
Demand curves for pharmacy-based rapid COVID-19 testing by county.

Considering each of the counties, 69.8% of private retail pharmacy clients in Nairobi, 68.3% in Siaya and 34.2% in Kisumu would be willing to pay KES 500 (US$ 4.57). However, if this price was doubled to KES 1,000 (US$ 9.15), approximately 50.0%, 27.5% and 15.6% would be willing to pay this price in Nairobi, Siaya and Kisumu counties, respectively.

Additionally, at these price and demand points, the demand for COVID-19 testing kits in Siaya (PED = 1.28) and Kisumu (PED = 1.12) was relatively price elastic, while demand in Nairobi was price inelastic (PED = 0.50). This means that the same change in price will result in a larger decrease in the number of clients willing to pay in Siaya and Kisumu counties than in Nairobi County.

### 3.4 Mean and Median maximum willingness to pay levels

**Table 4** presents the mean and median maximum prices that private retail pharmacy clients are willing to pay for COVID-19 rapid testing across selected sociodemographic factors. On average, the maximum willingness to pay price for a rapid COVID-19 test in a private retail pharmacy in Siaya was KES 197 (US$ 1.80) (95% CI: 42 – 351) higher compared to that in Kisumu (KES 445 (US$ 4.07) (95% CI: 371 – 519, p=0.013). Although lower than the average maximum WTP price in Siaya, the maximum WTP price in Nairobi was KES 176 (US$ 1.61) (95% CI: 73 – 280) higher than that in Kisumu (p=0.001). The maximum WTP price also varied by gender, level of education and income level. For instance, male clients were willing on average to pay a maximum WTP price of KES 182 (US$ 1.66) (95% CI: 73 – 291) higher compared to female clients (Max WTP = KES 452 (US$ 4.13) (95% CI: 389 – 516 p=0.001). Clients coming from households reporting an income above the minimum monthly wage of KES 13,572 (US$ 124.15) had a higher maximum WTP compared to their counterparts reporting less than the minimum monthly wage (p<0.001). Furthermore, the maximum WTP price increased with a client’s interest in getting COVID-19 at a pharmacy when in need of testing.

**Table 4:** Distribution of clients’ mean and median maximum willingness to pay by sociodemographic factors.

|  | Mean |  |  |  | Median |  |  |  |
| --- | --- | --- | --- | --- | --- | --- | --- | --- |
| | KES | US\$ | 95% CI (KES) | | KES | US\$ | IQR | |
|  |  |  | Lower | Upper |  |  | Lower | Upper |
| **Overall** | 552 | 5.05 | 497 | 607 | 500 | 4.57 | 200 | 750 |
| **County** |  |  |  |  |  |  |  |  |
| Nairobi | 621 | 5.68 | 550 | 692 | 500 | 4.57 | 500 | 1000 |
| Siaya | 641 | 5.86 | 483 | 799 | 500 | 4.57 | 200 | 750 |
| Kisumu | 445 | 4.07 | 372 | 518 | 300 | 2.74 | 100 | 500 |
| **Gender** |  |  |  |  |  |  |  |  |
| Female | 452 | 4.13 | 389 | 515 | 375 | 3.43 | 200 | 500 |
| Male | 634 | 5.80 | 550 | 719 | 500 | 4.57 | 200 | 1000 |
| **Current marital status** |  |  |  |  |  |  |  |  |
| Not married | 538 | 4.92 | 461 | 614 | 500 | 4.57 | 200 | 750 |
| Married | 559 | 5.11 | 486 | 632 | 500 | 4.57 | 200 | 700 |
| **Age group (years)** |  |  |  |  |  | - |  |  |
| 18-25 | 542 | 4.96 | 431 | 652 | 500 | 4.57 | 200 | 500 |
| 26-35 | 505 | 4.62 | 432 | 578 | 500 | 4.57 | 200 | 500 |
| 36-55 | 613 | 5.61 | 491 | 734 | 500 | 4.57 | 200 | 875 |
| 56+ | 584 | 5.34 | 389 | 778 | 500 | 4.57 | 200 | 1000 |
| **Level of education** |  |  |  |  |  |  |  |  |
| No Education | 286 | 2.62 | 140 | 432 | 200 | 1.83 | 100 | 300 |
| Primary education | 542 | 4.96 | 393 | 691 | 500 | 4.57 | 200 | 500 |
| Secondary education | 451 | 4.13 | 386 | 516 | 500 | 4.57 | 200 | 500 |
| Tertiary education | 678 | 6.20 | 589 | 766 | 500 | 4.57 | 300 | 1000 |
| **Employment status** |  |  |  |  |  |  |  |  |
| Not Employed | 571 | 5.22 | 465 | 677 | 500 | 4.57 | 200 | 750 |
| Employed | 539 | 4.93 | 476 | 603 | 500 | 4.57 | 200 | 600 |
| **Household income group** |  |  |  |  |  |  |  |  |
| Below monthly minimum wage (13,572) | 436 | 3.99 | 360 | 511 | 300 | 2.74 | 100 | 500 |
| Above monthly minimum wage | 678 | 6.20 | 602 | 754 | 500 | 4.57 | 300 | 1000 |
| **Interest in COVID-19 test at Pharmacy if in need** |  |  |  |  |  |  |  |  |
| Not at all interested | 282 | 2.58 | 87 | 478 | 200 | 1.83 | 50 | 200 |
| Somewhat interested | 525 | 4.80 | 405 | 645 | 500 | 4.57 | 200 | 750 |
| Very interested | 575 | 5.26 | 511 | 639 | 500 | 4.57 | 200 | 750 |

### 3.5 Determinants of the maximum willingness to pay

**Table 5** presents the predictors of the maximum willingness to pay for rapid COVID-19 testing. In a multivariable model accounting for age, gender, marital status, employment status, county, interest in COVID-19 testing at a pharmacy, and household income, factors that were positively correlated with a higher maximum WTP price included tertiary education, being from Siaya, having an interest in COVID-19 test and higher household income group. Being from Kisumu was negatively associated with the reported maximum WTP price.

**Table 5:**
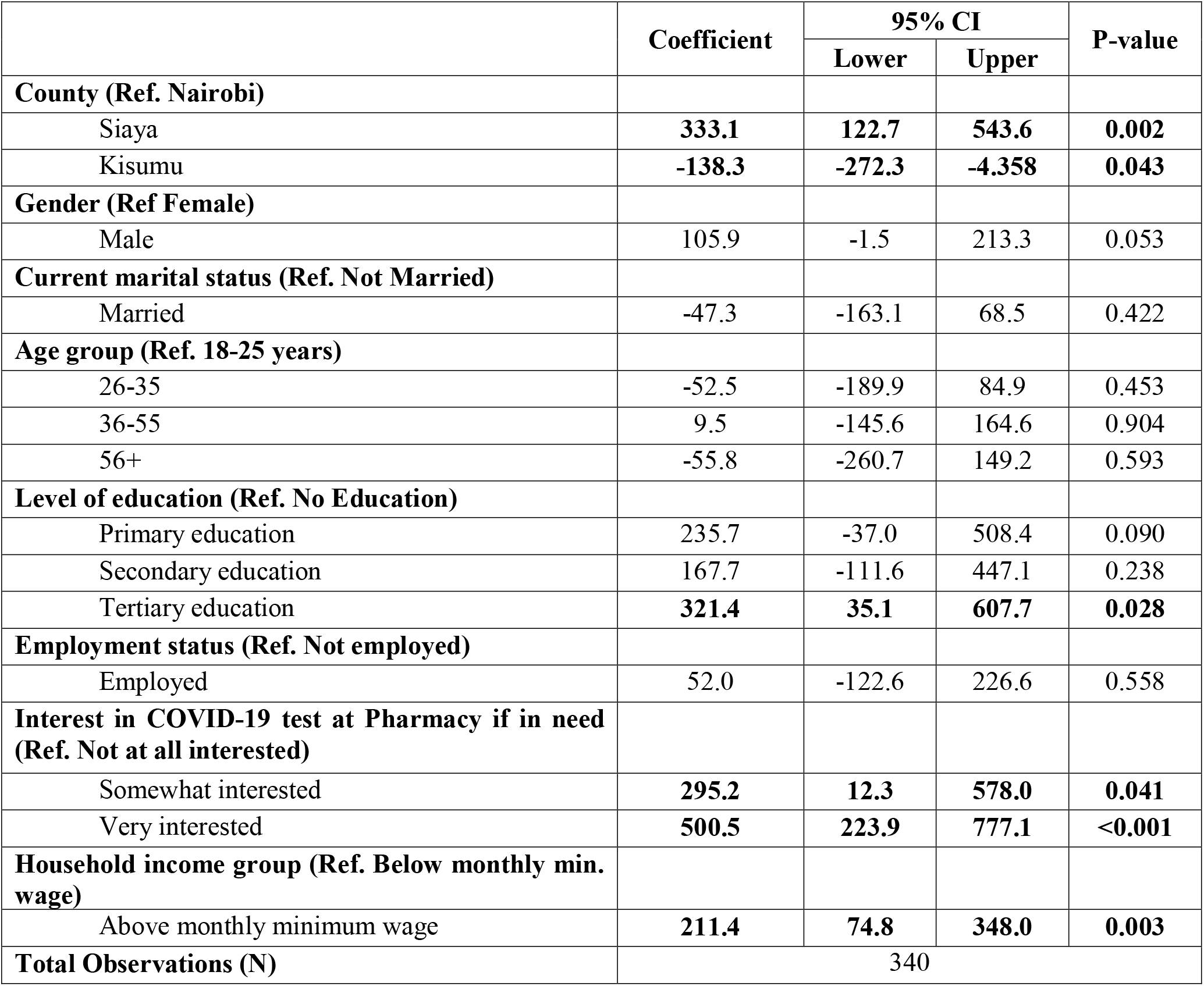
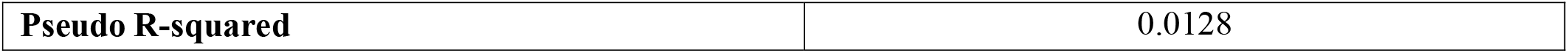
Determinants of the maximum willingness to pay for a rapid COVID-19 test at a private retail pharmacy.

|  | Coefficient | 95% CI |  | P-value |
| --- | --- | --- | --- | --- |
|  |  | Lower | Upper |  |
| **County (Ref. Nairobi)** |  |  |  |  |
| Siaya | **333.1** | **122.7** | **543.6** | **0.002** |
| Kisumu | **-138.3** | **-272.3** | **-4.358** | **0.043** |
| **Gender (Ref Female)** |  |  |  |  |
| Male | 105.9 | -1.5 | 213.3 | 0.053 |
| **Current marital status (Ref. Not Married)** |  |  |  |  |
| Married | -47.3 | -163.1 | 68.5 | 0.422 |
| **Age group (Ref. 18-25 years)** |  |  |  |  |
| 26-35 | -52.5 | -189.9 | 84.9 | 0.453 |
| 36-55 | 9.5 | -145.6 | 164.6 | 0.904 |
| 56+ | -55.8 | -260.7 | 149.2 | 0.593 |
| **Level of education (Ref. No Education)** |  |  |  |  |
| Primary education | 235.7 | -37.0 | 508.4 | 0.090 |
| Secondary education | 167.7 | -111.6 | 447.1 | 0.238 |
| Tertiary education | **321.4** | **35.1** | **607.7** | **0.028** |
| **Employment status (Ref. Not employed)** |  |  |  |  |
| Employed | 52.0 | -122.6 | 226.6 | 0.558 |
| **Interest in COVID-19 test at Pharmacy if in need (Ref. Not at all interested)** |  |  |  |  |
| Somewhat interested | **295.2** | **12.3** | **578.0** | **0.041** |
| Very interested | **500.5** | **223.9** | **777.1** | **<0.001** |
| **Household income group (Ref. Below monthly min. wage)** |  |  |  |  |
| Above monthly minimum wage | **211.4** | **74.8** | **348.0** | **0.003** |
| **Total Observations (N)** | 340 |  |  |  |

|  |  |
| --- | --- |
| **Pseudo R-squared** | 0.0128 |

## 4.0 DISCUSSION

This study examined the level and determinants of willingness to pay for rapid COVID-19 testing delivered through private retail pharmacies in Kenya. First, we found that respondents were willing to pay a mean price of KES 611 (US$ 5.59) and a median price of KES 506 (US$ 4.63). To the best of our knowledge, this is the first study in Africa to examine the level and determinants of willingness to pay for COVID-19 testing. Although no previous study has assessed the WTP for rapid diagnostic testing for COVID-19, a study conducted in Latin America reported a WTP of USD 45 (approximately KES 4,919.40; 1 USD ≈ KES 109.32) for laboratory COVID-19 PCR testing [42]. The mean and median WTP prices in our study were nearly 3 times lower than the cost of a laboratory COVID-19 test (KES 1,816.62) currently in use in Kenya [9]. This means that there is a high likelihood of uptake of pharmacy-based rapid COVID-19 testing if priced at between KES 500 and KES 1000.The finding that over 90% of the respondents indicated some interest in obtaining the test at a private retail pharmacy when in need further underscores the high likelihood for uptake.

Second, our analysis also found that the WTP price varied across study counties, probably reflective of rural-urban differences and corresponding differences in purchasing power. The variation in the mean maximum WTP may also reflect regional differences in perceived utility of COVID-19 testing delivered through pharmacies. This difference in WTP price could indicate the need for county-specific price-setting for pharmacy-based COVID-19 testing.

Third, household income was positively associated with the WTP for a COVID-19 test. A possible explanation for this includes the fact household income determines the disposable income that influences the household’s ability to pay for tests. It is anticipated that individuals or households with a higher income would be more willing to pay than those with a low income given their higher disposable income [43]. These findings are similar to those of the study conducted in Latin America [42].

Fourth, interestingly, clients’ interest in a COVID-19 rapid test was associated with WTP. This may have implications for designing strategies to increase uptake of COVID-19 testing across pharmacies. For instance, strategies can be tailored to enhance the population interest such as through raising awareness about rapid diagnostic testing through pharmacies, hence, the uptake of testing. Future studies can explore this further.

Findings from this study should be interpreted in light of some limitations. First, since the study was conducted in only 3 predominantly urban counties, the findings may not be generalizable to all counties in Kenya or other African country settings. Future studies can increase both the sample size and counties engaged to get further details about the variation in prices and obtain a more generalizable estimate. Second, as with all other stated preference elicitation approaches, WTP estimates from a contingent valuation method (a type of a stated preference elicitation method) are prone to hypothetical bias where participants may be inclined to state a higher willingness to pay price than they would pay in reality. However, a clear description of the settings and conducting the study during the COVID-19 pandemic hopefully makes our estimates realistic. Third, the linear estimation of the demand curve may not be entirely accurate as the relationship between quantity demanded and price may not be as linear as it is often a non-linear curve with a negative slope [44]. Future studies could include more starting bids to enable better plotting of the demand curve.

## 5.0 CONCLUSION

With the ongoing COVID-19 pandemic and low acquisition and uptake of COVID-19 vaccination in Kenya, testing remains a crucial response strategy. Private retail pharmacies can be leveraged to extend COVID-19 testing coverage through the provision of rapid testing. Our findings on WTP levels and determinants provide information that can inform price setting, price differentiation, subsidization and other program design features geared towards promoting affordability, equity and uptake.

## Data Availability

All data produced in the present study are available upon reasonable request to the authors

## Notes

### Competing Interest Statement

The authors have declared no competing interest.

### Funding Statement

This study was funded by the Foundation for Innovative New Diagnostics.

### Author Declarations

We received ethical approval from the Kenya Medical Research Institute (KEMRI) Scientific and Ethics Review Unit (SERU) under SERU number SSC: 4115

## REFERENCES

1. Darab MG, Keshavarz K, Sadeghi E, Shahmohamadi J, Kavosi Z: The economic burden of coronavirus disease 2019 (COVID-19): evidence from Iran. BMC Health Services Research 2021, 21(1):1–7.

2. Maital S, Barzani E: The global economic impact of COVID-19: A summary of research. Samuel Neaman Institute for National Policy Research 2020, 2020:1–12.

3. Patrick G, Walker C, Oliver W: The global impact of COVID-19 and strategies for mitigation and suppression. WHO Collaborating Centre for Infectious Disease Modelling, MRC Centre for Global Infectious Disease Analysis: Abdul Latif Jameel Institute for Disease and Emergency Analytics, Imperial College London 2020.

4. Coronavirus cases and deaths [https://www.worldometers.info/coronavirus/]

5. Guleid F, Akech S, Oyier I, Barasa E: What is the role of testing in the COVID-19 pandemic? (Evidence brief 4). 2020.

6. Torres I, Sippy R, Sacoto F: Assessing critical gaps in COVID-19 testing capacity: the case of delayed results in Ecuador. BMC public health 2021, 21(1):1–8.

7. Vandenberg O, Martiny D, Rochas O, van Belkum A, Kozlakidis Z: Considerations for diagnostic COVID-19 tests. Nature Reviews Microbiology 2021, 19(3):171–183.

8. The Global Fund: Diagnostics Pillar explainer. In.; 2020.

9. Barasa E, Kairu A, Maritim M, Were V, Akech S, Mwangangi M: Examining unit costs for COVID-19 case management in Kenya. BMJ global health 2021, 6(4):e004159.

10. Foundation for Innovative New Diagnostics (FIND): Test Directory. In.; 2021.

11. Abuya TO, Mutemi W, Karisa B, Ochola SA, Fegan G, Marsh V: Use of over-the-counter malaria medicines in children and adults in three districts in Kenya: implications for private medicine retailer interventions. Malaria journal 2007, 6(1):1–10.

12. Bigogo G, Audi A, Aura B, Aol G, Breiman RF, Feikin DR: Health-seeking patterns among participants of population-based morbidity surveillance in rural western Kenya: implications for calculating disease rates. International journal of Infectious diseases 2010, 14(11):e967–e973.

13. Fonck K, Mwai C, Rakwar J, Kirui P, Ndinya-Achola JO, Temmerman M: Healthcare-seeking behavior and sexual behavior of patients with sexually transmitted diseases in Nairobi, Kenya. Sexually transmitted diseases 2001, 28(7):367–371.

14. Onwujekwe O, Hanson K, Uzochukwu B: Do poor people use poor quality providersã Evidence from the treatment of presumptive malaria in Nigeria. Tropical medicine & international health : TM & IH 2011, 16(9):1087–1098.

15. Barasa E, Kazungu J, Orangi S, Kabia E, Ogero M, Kasera K: Indirect health effects of the COVID-19 pandemic in Kenya: a mixed methods assessment. BMC Health Services Research 2021, 21(1):1–16.

16. Smith F: Private local pharmacies in low-and middle-income countries: a review of interventions to enhance their role in public health. Tropical Medicine & International Health 2009, 14(3):362–372.

17. Mugo PM, Micheni M, Shangala J, Hussein MH, Graham SM, Rinke de Wit TF, Sanders EJ: Uptake and acceptability of oral HIV self-testing among community pharmacy clients in Kenya: a feasibility study. PloS one 2017, 12(1):e0170868.

18. Ortblad KF, Mogere P, Roche S, Kamolloh K, Odoyo J, Irungu E, Mugo NR, Pintye J, Baeten JM, Bukusi E: Design of a care pathway for pharmacy-based PrEP delivery in Kenya: results from a collaborative stakeholder consultation. BMC health services research 2020, 20(1):1–9.

19. Chiu C, Hunter LA, McCoy SI, Mfaume R, Njau P, Liu JX: Sales and pricing decisions for HIV self-test kits among local drug shops in Tanzania: a prospective cohort study. BMC health services research 2021, 21(1):1–11.

20. Cohen J, Fink G, Maloney K, Berg K, Jordan M, Svoronos T, Aber F, Dickens W: Introducing rapid diagnostic tests for malaria to drug shops in Uganda: a cluster-randomized controlled trial. Bulletin of the World Health Organization 2015, 93:142–151.

21. Cerda AA, García LY: Willingness to Pay for a COVID-19 Vaccine. Applied health economics and health policy 2021, 19(3):343–351.

22. Wong LP, Alias H, Wong P-F, Lee HY, AbuBakar S: The use of the health belief model to assess predictors of intent to receive the COVID-19 vaccine and willingness to pay. Human vaccines & immunotherapeutics 2020, 16(9):2204–2214.

23. García LY, Cerda AA: Contingent assessment of the COVID-19 vaccine. Vaccine 2020, 38(34):5424–5429.

24. Ashburn K, Antelman G, N’Goran MK, Jahanpour O, Yemaneberhan A, N’Guessan Kouakou B, Kazemi E, Duffy M, Adama P, Kajoka D: Willingness to use HIV self-test kits and willingness to pay among urban antenatal clients in Cote d’Ivoire and Tanzania: a cross-sectional study. Tropical Medicine & International Health 2020, 25(9):1155–1165.

25. Thirumurthy H, Masters SH, Agot K: Willingness to pay for HIV self-tests among women in Kenya: implications for subsidy and pricing policies. Journal of acquired immune deficiency syndromes (1999) 2018, 78(2):e8.

26. PPB: Online Services Portal: License Status. In.: Pharmacy and Poisons Board; 2019.

27. Statista: Cumulative number of confirmed coronavirus (COVID-19) cases in Kenya by county. In.: Statista; 2021.

28. Worldometer: Kenya Coronavirus Cases. In.: Worldometer; 2021.

29. Cooper JC: Optimal bid selection for dichotomous choice contingent valuation surveys. Journal of Environmental Economics and Management 1993, 24(1):25–40.

30. Vaughan WJ, Darling AH: The Optimal Sample Size for Contingent Valuation Surveys. In.: Working Paper No. ENV-136. April. Washington, DC: Inter-American Development …; 2000.

31. Aizaki H, Nakatani T, Sato K: Stated preference methods using R: CRC Press; 2014.

32. Hanemann M, Loomis J, Kanninen B: Statistical efficiency of double-bounded dichotomous choice contingent valuation. American journal of agricultural economics 1991, 73(4):1255–1263.

33. Calia P, Strazzera E: Bias and efficiency of single vs double bound models for contingent valuation studies: A Monte Carlo analysis. Fondazione Eni Enrico Mattei Working Paper 1999(10.99).

34. Palmieri N, Suardi A, Pari L: Italian consumers’ willingness to pay for eucalyptus firewood. Sustainability 2020, 12(7):2629.

35. Wright A: REDCap: A tool for the electronic capture of research data. Journal of Electronic Resources in Medical Libraries 2016, 13(4):197–201.

36. Boyle KJ, Bishop RC, Welsh MP: Starting point bias in contingent valuation bidding games. Land economics 1985, 61(2):188–194.

37. Yoo S-H, Shin E: Analyzing Dichotomous Choice Contingent Valuation Data with Zero Observations: A Mixture Model. Korean Economic Review 2001, 17(2):311–328.

38. Hanemann WM: Welfare evaluations in contingent valuation experiments with discrete responses. American journal of agricultural economics 1984, 66(3):332–341.

39. Ringel JS, Hosek SD, Vollaard BA, Mahnovski S: The elasticity of demand for health care. A review of the literature and its application to the military health system 2002.

40. Kim TK: T test as a parametric statistic. Korean journal of anesthesiology 2015, 68(6):540.

41. Sigelman L, Zeng L: Analyzing censored and sample-selected data with Tobit and Heckit models. Political analysis 1999, 8(2):167–182.

42. Trudeau JM, Alicea-Planas J, Vásquez WF: The value of COVID-19 tests in Latin America. Economics & Human Biology 2020, 39:100931.

43. Organization WH: The impact of health expenditure on households and options for alternative financing. In.; 2004.

44. Feldman R, Dowd B: What does the demand curve for medical care measure? Journal of Health Economics 1993, 12(2):193–200.

