## Appendix 1 for "Level and determinants of willingness to pay for rapid COVID-19 testing delivered through private retail pharmacies in Kenya"

**APPENDIX 1 – WILLINGNESS TO PAY (WTP) QUESTIONNAIRE**

**BASIC INFORMATION**

1. Questionnaire Number: [__] [	] [	] [	]
2. County Name: ____________________________________________________
3. Interviewer Name: _________________________________________________
4. Interview Date: Mo. ___ Day___ Yr. ___
5. Time Interview Began: ______________
6. Time Interview Completed: ______________

**SOCIODEMOGRAPHIC CHARACTERISTICS**

| QN | QUESTION | RESPONSE | CODE | SKIP |
| --- | --- | --- | --- | --- |
| 100 | How old were you at your last birthday? | YEARS __________ |  |  |
| 101 | What is your gender? | Female  Male | 1  2 |  |
| 102 | What is your marital status? | Married/In-union  Widowed, Separated, Divorced  Single | 1  2  3 |  |
| 103 | What is your highest level of education?  PROBE FOR THE HIGHEST LEVEL COMPLETED | No education  Primary education  Secondary education  Tertiary (college or university) | 1  2  3  4 |  |
| 104 | Are you employed? | Yes  No | 1  2 | GO TO 104 |
| 105 | What sector are you employed in? | Formal sector  Informal sector | 1  2 |  |
| 106 | What is your family’s TOTAL INCOME per month? | KES__________________ |  |  |

**WILLINGNESS TO PAY**

**BLOCK 1**

| **QN** | **QUESTION** | **RESPONSE** | **CODE** | **SKIP** |
| --- | --- | --- | --- | --- |
| 200 | **READ TO CLIENT:**  I would like to ask you some questions about your willingness to pay for a new COVID-19 test to be obtained at pharmacies like this one. In answering these questions, please bear in mind the following:  1. Assume that your income will stay the same.  2. COVID-19 testing services are available in public health facilities as well as private laboratories. | |  |  |
| 201 | If the price of the COVID-19 testing kit at the Pharmacy was set at **KES 500**, would you be willing to pay this price? | Yes  No | 1  2 | GO TO 202  GO TO 203 |
| 202 | Suppose that instead of **KES 500,** the price of the COVID-19 testing kit at the Pharmacy was set at **KES 1,000**, would you be willing to pay this price? | Yes  No | 1  2 | GO TO 204  GO TO 204 |
| 203 | Suppose that instead of **KES 500**, the price of the COVID-19 testing kit at the Pharmacy was set at **KES 300**, would you be willing to pay this price? | Yes  No | 1  2 | GO TO 204  GO TO 204 |
| 204 | What is the most you would be willing to pay to get a COVID-19 test at a pharmacy? | KES__________________ |  |  |
| 205 | If the price of a COVID-19 testing kit at the pharmacy was beyond what you are willing and able to pay, what would you do? | Not test for COVID-19  Go somewhere else to test (hospital)  I don’t know | 1  2  99 | GO TO 206  GO TO 206  GO TO 206 |
| 206 | Would you say that you are interested in getting a COVID-19 test at the pharmacy if you ever thought you needed to get tested? | Not at all interested  Somewhat interested  Very interested | 1  2  3 |  |
| 207 | What service had you come to seek at the pharmacy? |  |  |  |

**BLOCK 2**

| QN | QUESTION | RESPONSE | CODE | SKIP |
| --- | --- | --- | --- | --- |
| 200 | READ TO CLIENT:  I would like to ask you some questions about your willingness to pay for a new COVID-19 test to be obtained at pharmacies like this one. In answering these questions, please bear in mind the following:  1. Assume that your income will stay the same.  2. COVID-19 testing services are available in public health facilities as well as private laboratories | |  |  |
| 201 | If the price of the COVID-19 testing kit at the Pharmacy was set at **KES 1,000,** would you be willing to pay this price? | Yes  No | 1  2 | GO TO 202  GO TO 203 |
| 202 | Suppose that instead of **KES 1,000**, the price of the COVID-19 testing kit at the Pharmacy was set at **KES** **1,500**, would you be willing to pay this price? | Yes  No | 1  2 | GO TO 204  GO TO 204 |
| 203 | Suppose that instead of **KES 1,000**, the price of the COVID-19 testing kit at the Pharmacy was set at **KES 750**, would you be willing to pay this price? | Yes  No | 1  2 | GO TO 204  GO TO 204 |
| 204 | What is the most you would be willing to pay to get a COVID-19 test at a pharmacy?  (Remind the respondent about the amount indicated as “Yes” above) | KES__________________ |  |  |
| 205 | If the price of a COVID-19 testing kit at the pharmacy was beyond what you are willing and able to pay, what would you do? | Not test for COVID-19  Go somewhere else to test (hospital)  I don’t know | 1  2  99 | GO TO 206  GO TO 206  GO TO 206 |
| 206 | Would you say that you are interested in getting a COVID-19 test at the pharmacy if you ever thought you needed to get tested? | Not at all interested  Somewhat interested  Very interested | 1  2  3 |  |
| 207 | What service had you come to seek at the pharmacy? |  |  |  |
